# The Link Between Atrial Fibrillation And Cognitive Impairment: A Systematic Review And Meta-Analysis

**DOI:** 10.1101/2023.08.30.23294787

**Authors:** Abhimanyu Agarwal, Mohamed A. Mostafa, Muhammad Imtiaz Ahmad, Elsayed Z. Soliman

**Affiliations:** Epidemiological Cardiology Research Center, Section on Cardiovascular Medicine, Department of Medicine, Wake Forest University School of Medicine, Winston-Salem, North Carolina; Department of Internal Medicine, Section on Hospital Medicine, Medical College of Wisconsin, Wauwatosa, Wisconsin

**Author notes:** Corresponding Author Abhimanyu Agarwal, MBBS Epidemiological Cardiology Research Center (EPICARE) Wake Forest School of Medicine Medical Center Blvd, Winston Salem, NC 27157.

**Keywords:** Keywords, AF, Cognitive Impairment, Dementia, Meta-analysis, Stroke

## Abstract

Atrial fibrillation (AF) affects a substantial population of individuals aged ≥55 years globally and has been associated with an increased risk of cognitive impairment (CImp) due to the shared risk factors such as aging, obesity, and diabetes. It is essential to comprehend their unique relationship to manage them effectively to reduce its impact on public health systems. We performed a PubMed search of English language papers published till 31 May 2023. Data was extracted and effect measures were calculated. Meta-analysis of 33 studies revealed a significant association between AF and CImp after adjusting for comorbidities, with considerable between-study heterogeneity (pooled OR: 1.62; 95% CI: 1.43, 1.81, I^2^ = 94.143). Prospective and cross-sectional studies revealed substantial associations. CImp in AF may be mediated through silent brain infarctions, microemboli, brain atrophy, and inflammation. Future research should prioritize investigating the mechanisms and etiology of this association, as well as potential treatment options.

## INTRODUCTION

Atrial fibrillation has been dubbed as the 21st-century cardiovascular disease epidemic^1–4^. With approximately 33 million people worldwide aged 55 and over affected by it, it is common among the elderly. AF is presently the most widely treated arrhythmia in clinical settings^5,6^ with $26 billion in healthcare costs related to its management and complications. Asia is predicted to have 72 million AF patients by 2050, with 2.9 million of them at risk for strokes linked to AF^7^. One in nine persons has subjective cognitive decline (SCD) and is more prevalent in people ≥ 65 years old compared to younger populations aged 45 to 64 (11.7% vs. 10.8%), respectively. Additionally, according to the Centers for Disease Control (CDC), 10.6% of women and 11.3% of men have SCD. It is crucial to remember that AF is associated with a 39% higher risk of cognitive impairment (CImp)^8^.

The shared risk factors between AF and CImp, such as aging, heart disease, obesity, diabetes, etc.^8,9^, which can occur alone or in combination^5^, underscore the need to investigate association between these risk factors and AF to develop preventive strategies. Studies demonstrate an increased risk of CImp and dementia associated with AF^10^, irrespective of a history of stroke^11,12^. Similar risk ratio estimates of 2.43^13^ and 2.70^11^ for CImp/dementia among people with AF were found in two meta-analyses. Persistent AF, but not paroxysmal AF, was associated with worse cognitive performance, according to an analysis of the Atherosclerosis Risk in Communities (ARIC) study^14^. These results imply that AF duration may have an additional effect on cognitive performance in addition to a mere presence of AF^15–18^.

There is significant consumption of Public health resources due to AF and its associated complications including CImp^19^. Worldwide, approximately 35.6 million people were estimated to have dementia in 2010, and it is predicted that this figure would roughly double every 20 years, reaching 65.7 million in 2030 and 115.4 million in 2050^20^. A comprehensive review that only considers studies involving individuals with no history of stroke to examine the association between AF and CImp is required to draw more conclusive findings. By establishing a relationship between AF and CImp independent of stroke or other CVD risk factors, it will highlight the urgent need to develop strategies to mitigate this significant relationship.

## METHODS

Following the Preferred Reporting Items for Systematic Reviews and Meta-Analyses (PRISMA)^21^ standards, this systematic review and meta-analysis were conducted.

### Data Sources

PubMed database was systematically searched from inception until 31 May 2023 using the search terms “Atrial Fibrillation and Cognitive Decline”, “Atrial Fibrillation and Cognitive Impairment”, “Atrial Fibrillation and Cognitive Function”, and “Atrial Fibrillation and Dementia” were utilized to retrieve studies involving individuals with cognitive impairment. Additionally, the references of the included studies and pertinent reviews were screened to identify any additional eligible studies. In total, 2892 studies were included in the search process.

### Study Selection

This systematic review included studies which met the following inclusion criteria:

1. Publication in the English language through May 2023.
2. Investigating the relationship between AF and CImp.
3. Reporting event rates or relative risks (RR)/odds ratios (OR) or hazard ratios (HR) for comparison groups.
4. Encompassing all observational study designs (cohort, cross-sectional, prospective, retrospective).
5. Studies in which cognitive assessment was done using standardized assessment scales eg Mini Mental State Examination (MMSE), Montreal Cognitive Assessment (MoCA), Modified Mini-Mental State Examination (3MS), informant questionnaire on cognitive decline in the elderly (IQCODE), Delayed Word Recall Test (DWRT), Wechsler Adult Intelligence Scale-Revised’s Digit Symbol Substitution Test (DSST), Word Fluency Test (WFT), etc.

Exclusion Criteria:

1. Studies with patients who had experienced stroke/ were suffering from vascular dementia/ underwent surgery/ had underlying psychological disorders/ cancer.
2. Patients taking antiarrhythmic medications.
3. Studies discussing pathophysiology of AF and dementia.
4. Did not report data about cognitive decline or only have brain imaging results/biomarkers/genetic markers.
5. Did not present original data, or were editorials, case reports, case series, systematic reviews/meta analysis.

Using the established criteria, we reviewers assessed the eligibility of studies included.

### Study Identification

We screened articles by title and abstract using predefined inclusion and exclusion criteria and a standardized data form. If the article did not meet inclusion criteria based on the abstract, the full text was not reviewed. The inclusion of full text was decided by consensus. All results were exported to Zotero, an open-source research tool for organizing and analyzing data, where duplicates were eliminated.

### Data Extraction and Outcomes

A structured data collection form was employed to gather information from each study, including study design, patient characteristics, baseline variables, cognitive evaluation method, follow-up duration in years, AF ascertainment. In cases where duplicate studies were found and studies reported the same outcome measure, only the larger study was included in the analysis.

### Quality Assessment

A consensus was reached to address any differences. The included studies’ epidemiological and clinical data were extracted using standardized forms. The Newcastle–Ottawa Scale (NOS) was employed to evaluate the quality of the article. NOS score ≥6 stars were defined as high quality, and NOS score <6 stars as low quality.

### Data Analysis

We extracted study baseline characteristics, event rates and sample sizes and calculated odds ratio for the primary and secondary outcomes using JASP 0.17.2.1 and the Campbell Effect Size Calculator for all statistical calculations. Statistics were deemed significant at a p-value of 0.05 level of significance.

When both CImp and dementia associations were separately reported, the outcome for the entire population was considered, as dementia is a subpopulation of CImp. We utilized meta-regression analysis and subgroup analysis (comparing cross-sectional and cohort studies) to explore potential sources of variance across studies. Age, hypertension, coronary artery disease, diabetes mellitus, and hyperlipidemia were all taken into account during the meta-regression analysis. Several sensitivity assessments were also conducted to investigate AF’s association with CImp/dementia beyond stroke.

A random-effects model was used to account for variability within and across studies. The Higgins I-squared statistic (I^2^) was used to determine the degree of heterogeneity. The risk of publication bias was shown using funnel plots and quantified using Egger’s regression test. To display the impact magnitude in each research and the combined estimate, a forest plot was used. A subgroup analysis was carried out to look at how the study design (cohort vs. cross-sectional) affected the result.

## RESULTS

This meta-analysis included 33 studies^12,18,22–52^ in total, and Figure 1 shows a thorough flow diagram of the method used to choose the studies. Table 1 shows characteristics of the sample population included. Of these investigations, two were case-control studies, four were retrospective in nature, 23 had a prospective design, and four provided cross-sectional analyses. Between 57 and 665,330 people were enrolled in the studies that were considered. Electrocardiogram (ECG) and the MMSE were the two tests that were used the most commonly to diagnose AF and CImp, respectively. The funnel plot was examined visually, and Egger’s test (p = 0.296) revealed that there was no publication bias.

**Figure 1:**
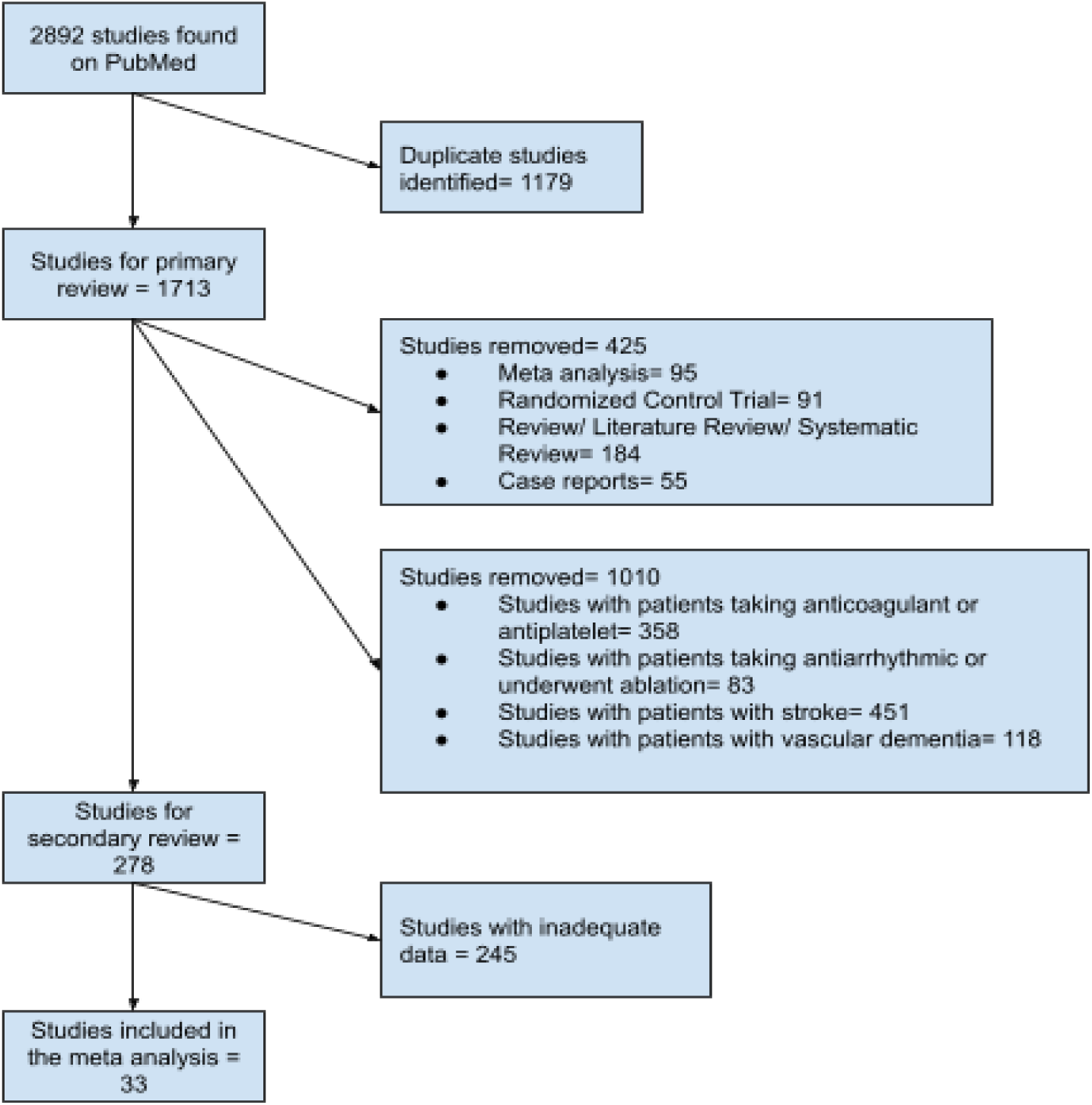
Study Selection Using Prisma Technique

**Table 1:**
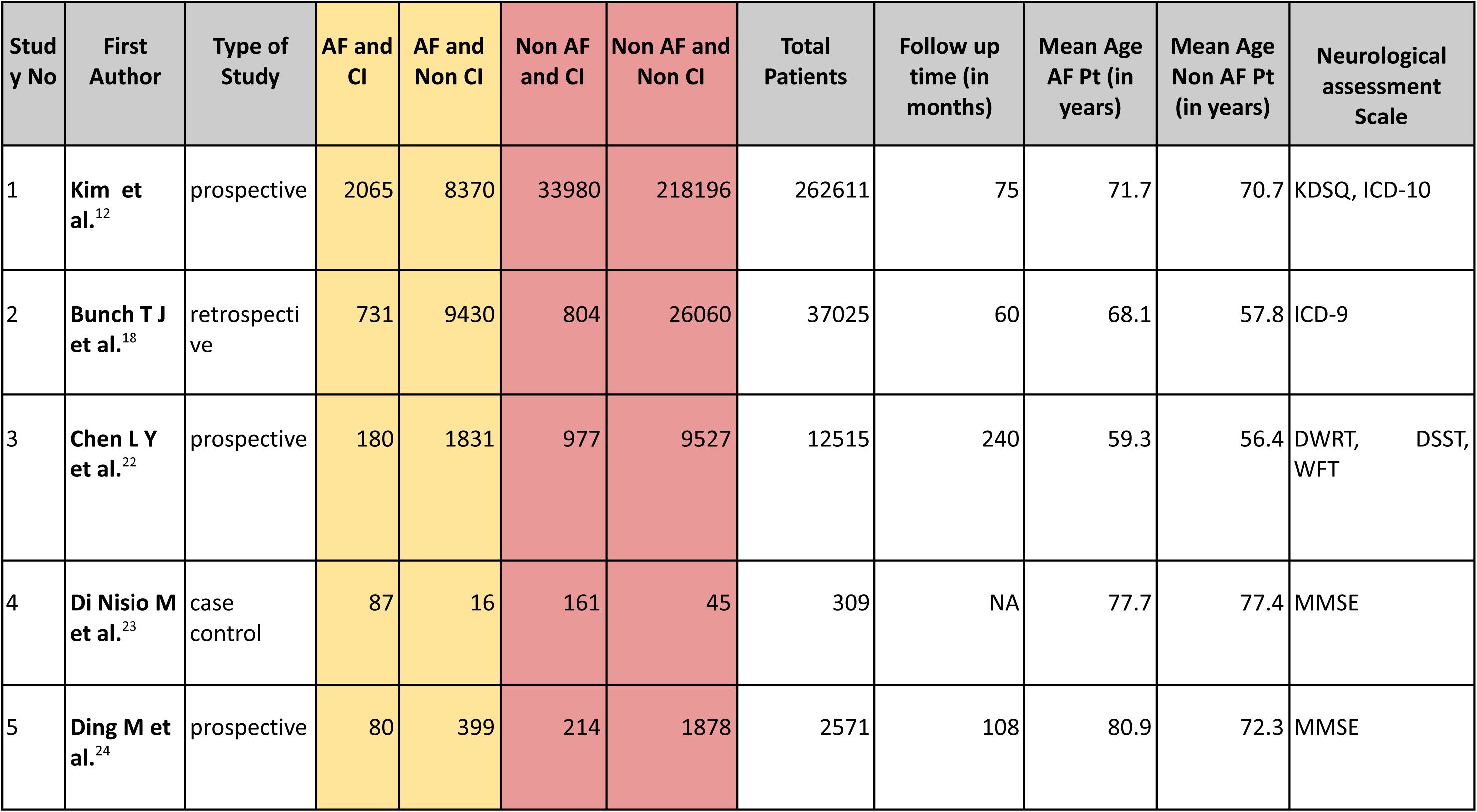
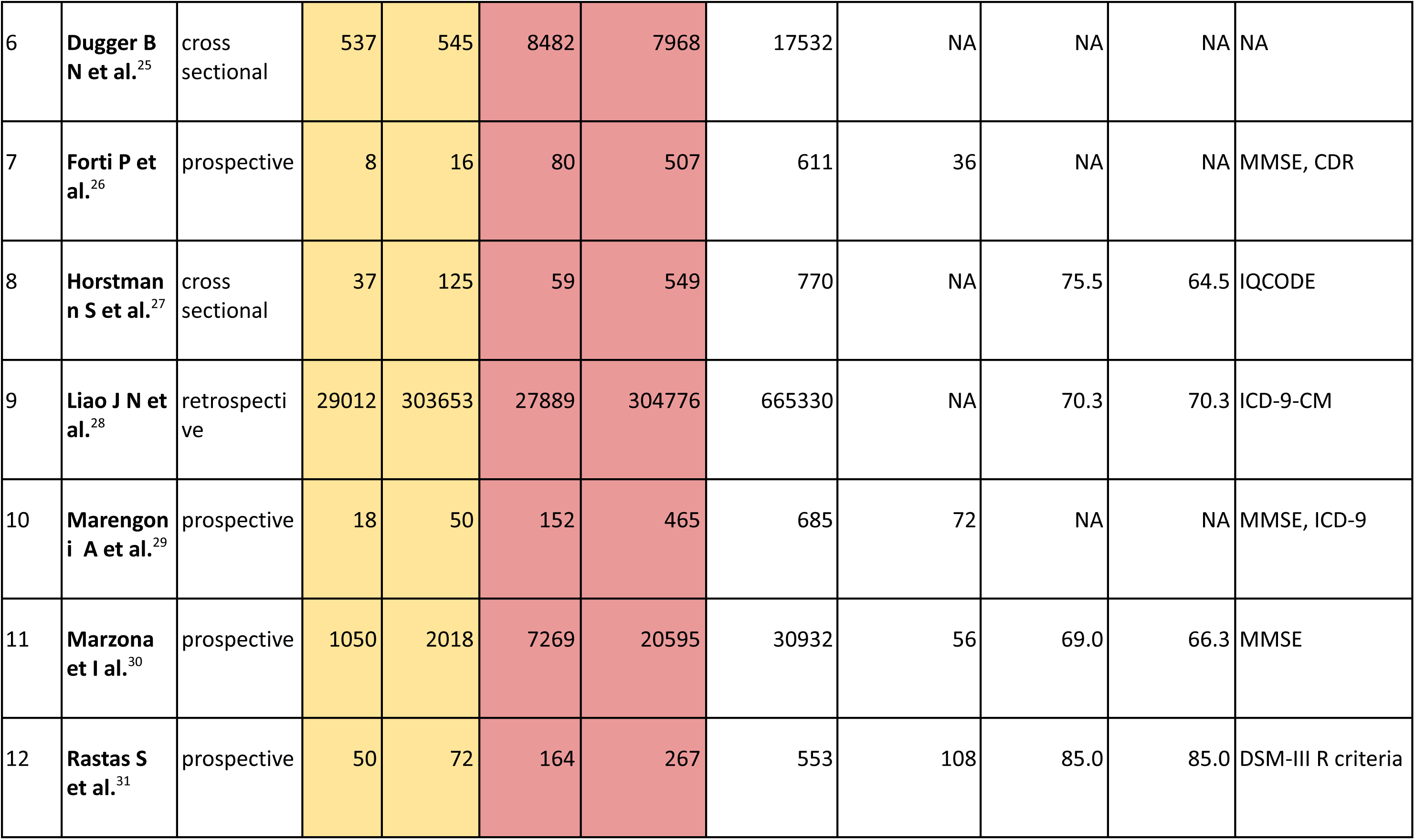
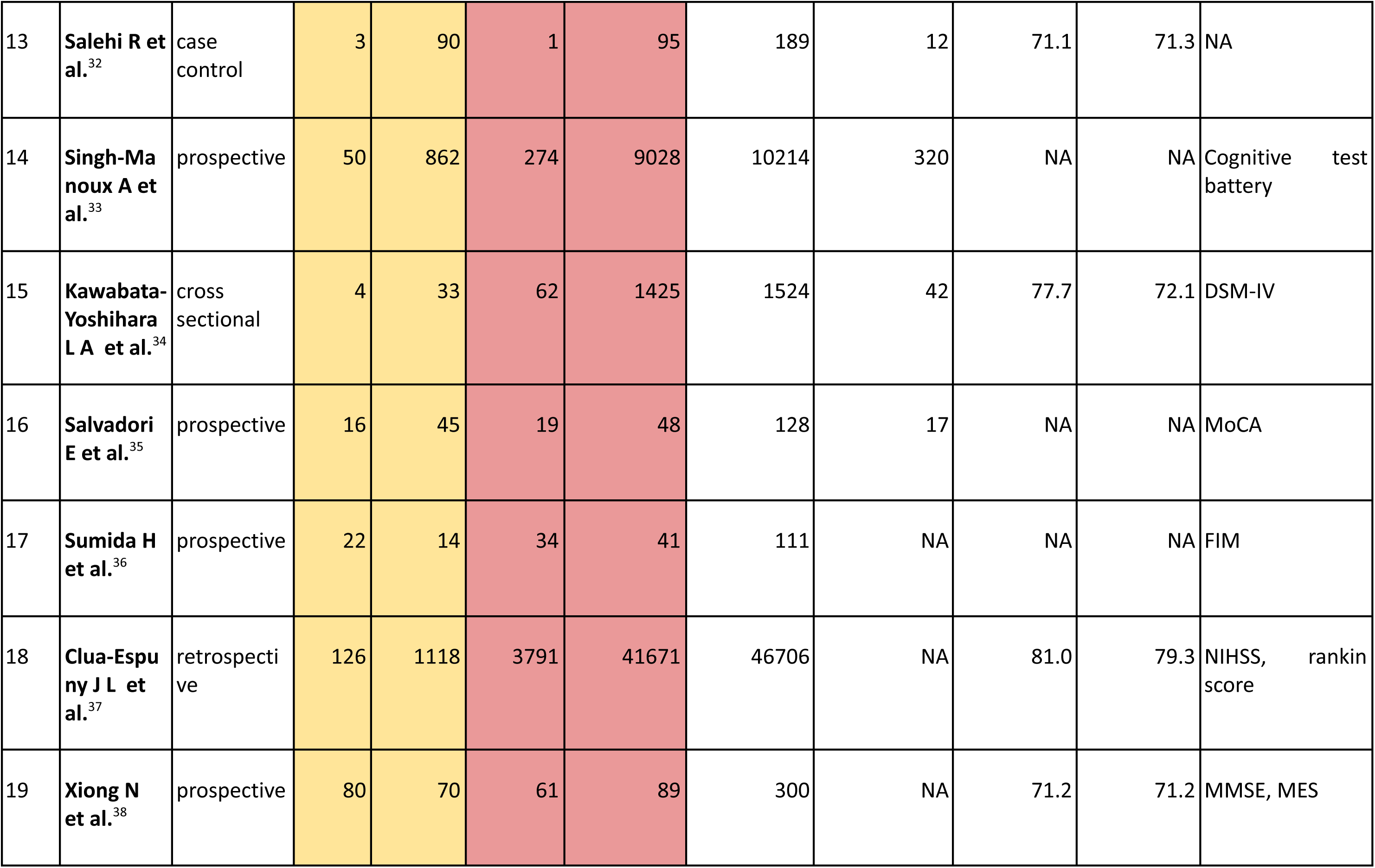
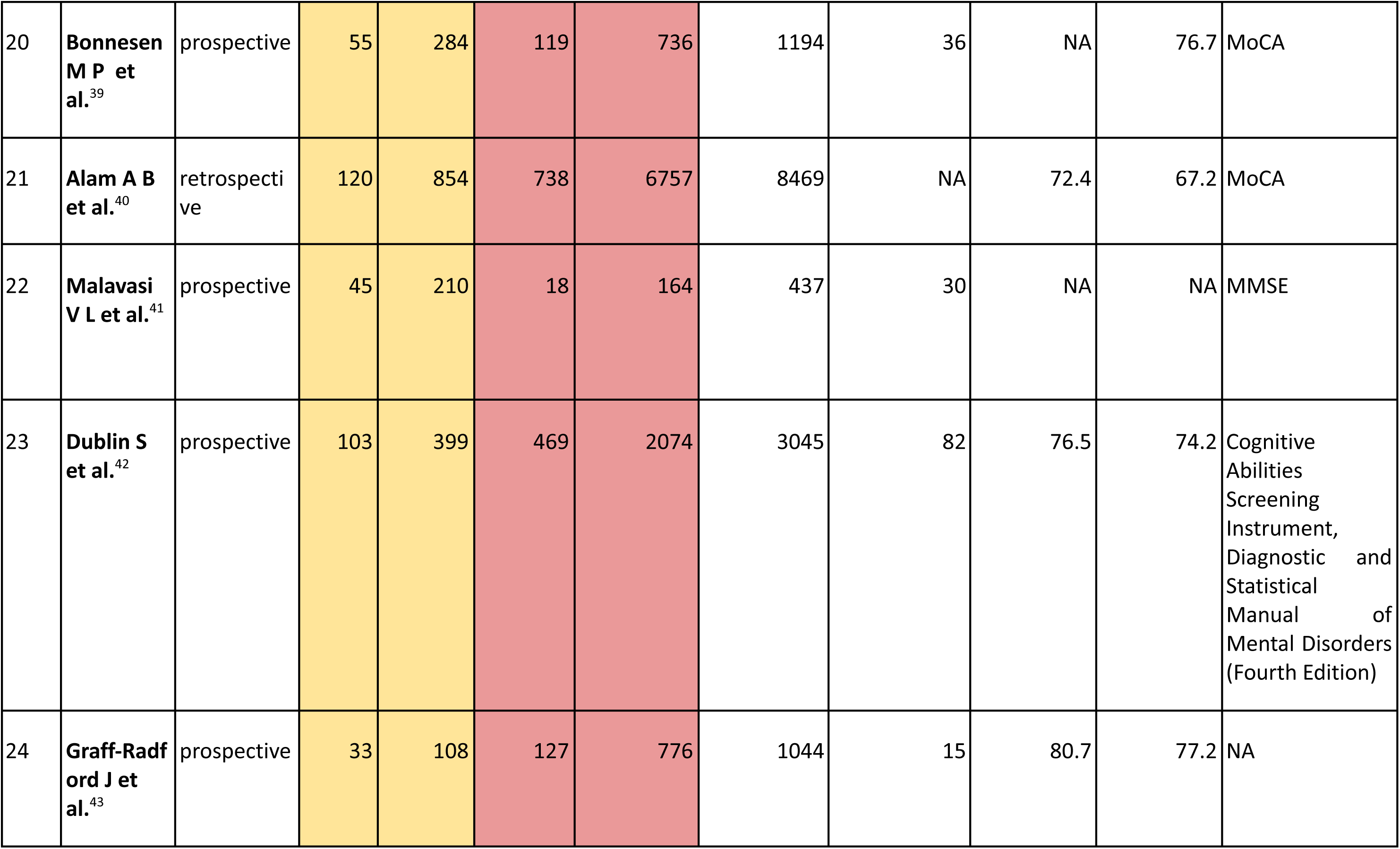
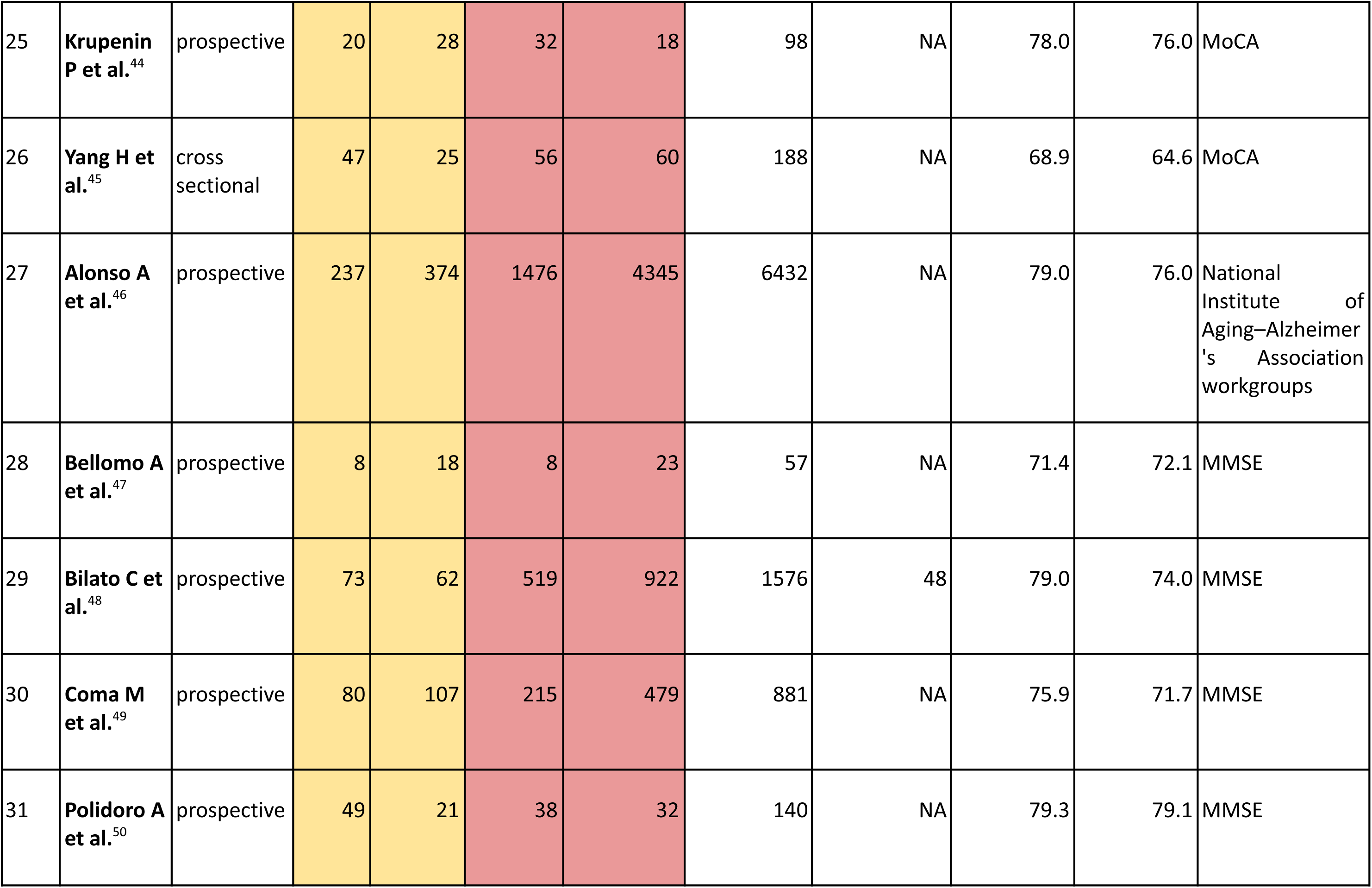
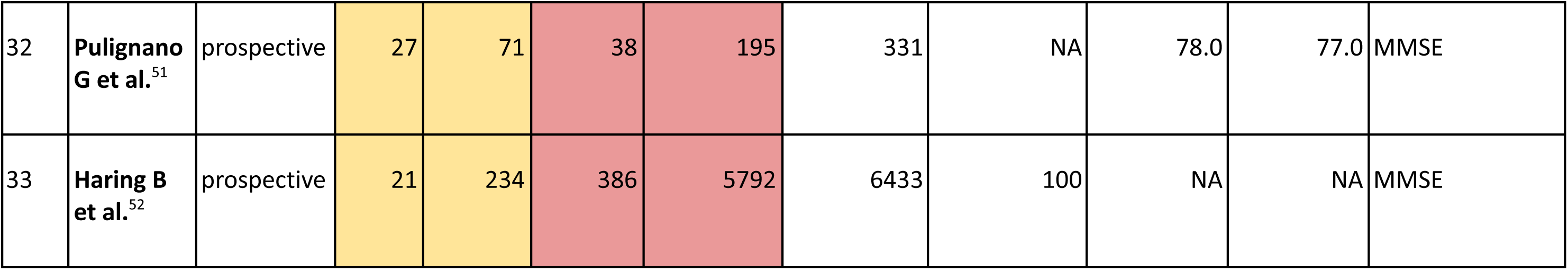
Basic Characteristics Of Study Population

| Study No | First Author | Type of Study | AF and CI | AF and Non CI | Non AF and CI | Non AF and Non CI | Total Patients | Follow up time (in months) | Mean Age AF Pt (in years) | Mean Age Non AF Pt (in years) | Neurological assessment Scale |
| --- | --- | --- | --- | --- | --- | --- | --- | --- | --- | --- | --- |
| 1 | Kim et al.<sup>12</sup> | prospective | 2065 | 8370 | 33980 | 218196 | 262611 | 75 | 71.7 | 70.7 | KDSQ, ICD-10 |
| 2 | Bunch T J et al.<sup>18</sup> | retrospective | 731 | 9430 | 804 | 26060 | 37025 | 60 | 68.1 | 57.8 | ICD-9 |
| 3 | Chen L Y et al.<sup>22</sup> | prospective | 180 | 1831 | 977 | 9527 | 12515 | 240 | 59.3 | 56.4 | DWRT, DSST, WFT |
| 4 | Di Nisio M et al.<sup>23</sup> | case control | 87 | 16 | 161 | 45 | 309 | NA | 77.7 | 77.4 | MMSE |
| 5 | Ding M et al.<sup>24</sup> | prospective | 80 | 399 | 214 | 1878 | 2571 | 108 | 80.9 | 72.3 | MMSE |
| 6 | **Dugger B N et al.**<sup>25</sup> | cross sectional | 537 | 545 | 8482 | 7968 | 17532 | NA | NA | NA | NA |
| 7 | **Forti P et al.**<sup>26</sup> | prospective | 8 | 16 | 80 | 507 | 611 | 36 | NA | NA | MMSE, CDR |
| 8 | **Horstman n S et al.**<sup>27</sup> | cross sectional | 37 | 125 | 59 | 549 | 770 | NA | 75.5 | 64.5 | IQCODE |
| 9 | **Liao J N et al.**<sup>28</sup> | retrospective | 29012 | 303653 | 27889 | 304776 | 665330 | NA | 70.3 | 70.3 | ICD-9-CM |
| 10 | **Marengoni A et al.**<sup>29</sup> | prospective | 18 | 50 | 152 | 465 | 685 | 72 | NA | NA | MMSE, ICD-9 |
| 11 | **Marzona et al.**<sup>30</sup> | prospective | 1050 | 2018 | 7269 | 20595 | 30932 | 56 | 69.0 | 66.3 | MMSE |
| 12 | **Rastas S et al.**<sup>31</sup> | prospective | 50 | 72 | 164 | 267 | 553 | 108 | 85.0 | 85.0 | DSM-III R criteria |
| 13 | **Salehi R et al.**<sup>32</sup> | case control | 3 | 90 | 1 | 95 | 189 | 12 | 71.1 | 71.3 | NA |
| 14 | **Singh-Ma**<br>**noux A et al.**<sup>33</sup> | prospective | 50 | 862 | 274 | 9028 | 10214 | 320 | NA | NA | Cognitive test<br>battery |
| 15 | **Kawabata-**<br>**Yoshihara**<br>**L A et al.**<sup>34</sup> | cross<br>sectional | 4 | 33 | 62 | 1425 | 1524 | 42 | 77.7 | 72.1 | DSM-IV |
| 16 | **Salvadori**<br>**E et al.**<sup>35</sup> | prospective | 16 | 45 | 19 | 48 | 128 | 17 | NA | NA | MoCA |
| 17 | **Sumida H**<br>**et al.**<sup>36</sup> | prospective | 22 | 14 | 34 | 41 | 111 | NA | NA | NA | FIM |
| 18 | **Clua-Espu**<br>**ny J L et al.**<sup>37</sup> | retrospecti<br>ve | 126 | 1118 | 3791 | 41671 | 46706 | NA | 81.0 | 79.3 | NIHSS, rankin<br>score |
| 19 | **Xiong N**<br>**et al.**<sup>38</sup> | prospective | 80 | 70 | 61 | 89 | 300 | NA | 71.2 | 71.2 | MMSE, MES |
| 20 | **Bonnesen M P et al.**<sup>39</sup> | prospective | 55 | 284 | 119 | 736 | 1194 | 36 | NA | 76.7 | MoCA |
| 21 | **Alam A B et al.**<sup>40</sup> | retrospective | 120 | 854 | 738 | 6757 | 8469 | NA | 72.4 | 67.2 | MoCA |
| 22 | **Malavasi V L et al.**<sup>41</sup> | prospective | 45 | 210 | 18 | 164 | 437 | 30 | NA | NA | MMSE |
| 23 | **Dublin S et al.**<sup>42</sup> | prospective | 103 | 399 | 469 | 2074 | 3045 | 82 | 76.5 | 74.2 | Cognitive Abilities Screening Instrument, Diagnostic and Statistical Manual of Mental Disorders (Fourth Edition) |
| 24 | **Graff-Radford J et al.**<sup>43</sup> | prospective | 33 | 108 | 127 | 776 | 1044 | 15 | 80.7 | 77.2 | NA |
| 25 | **Krupenin P et al.**<sup>44</sup> | prospective | 20 | 28 | 32 | 18 | 98 | NA | 78.0 | 76.0 | MoCA |
| 26 | **Yang H et al.**<sup>45</sup> | cross sectional | 47 | 25 | 56 | 60 | 188 | NA | 68.9 | 64.6 | MoCA |
| 27 | **Alonso A et al.**<sup>46</sup> | prospective | 237 | 374 | 1476 | 4345 | 6432 | NA | 79.0 | 76.0 | National Institute of Aging–Alzheimer's Association workgroups |
| 28 | **Bellomo A et al.**<sup>47</sup> | prospective | 8 | 18 | 8 | 23 | 57 | NA | 71.4 | 72.1 | MMSE |
| 29 | **Bilato C et al.**<sup>48</sup> | prospective | 73 | 62 | 519 | 922 | 1576 | 48 | 79.0 | 74.0 | MMSE |
| 30 | **Coma M et al.**<sup>49</sup> | prospective | 80 | 107 | 215 | 479 | 881 | NA | 75.9 | 71.7 | MMSE |
| 31 | **Polidoro A et al.**<sup>50</sup> | prospective | 49 | 21 | 38 | 32 | 140 | NA | 79.3 | 79.1 | MMSE |
| 32 | **Pulignano G et al.**<sup>51</sup> | prospective | 27 | 71 | 38 | 195 | 331 | NA | 78.0 | 77.0 | MMSE |
| 33 | **Haring B et al.**<sup>52</sup> | prospective | 21 | 234 | 386 | 5792 | 6433 | 100 | NA | NA | MMSE |

### Analysis of Individual Results

The analysis involved 33 studies^12,18,22–52^ in total, and the pooled analysis showed a significant correlation between AF and an elevated risk of outcome of interest i.e. CImp (pooled OR: 1.62; 95% CI: 1.43, 1.81; df: 32) with significant between-study heterogeneity (I^2^ = 94.143, 𝛕^2^ = 0.080, 𝛕 = 0.283, h^2^ = 17.072) (Figure 2, 3). The cook’s distance examined the influence measurements and found six outlier studies^18,23,30,34,39,47^. The between-study heterogeneity was marginally reduced once these outliers were taken into account (27 studies; pooled OR: 1.58; 95% CI: 1.38, 1.79; df: 26; I^2^ = 93.320).

**Figure 2:**
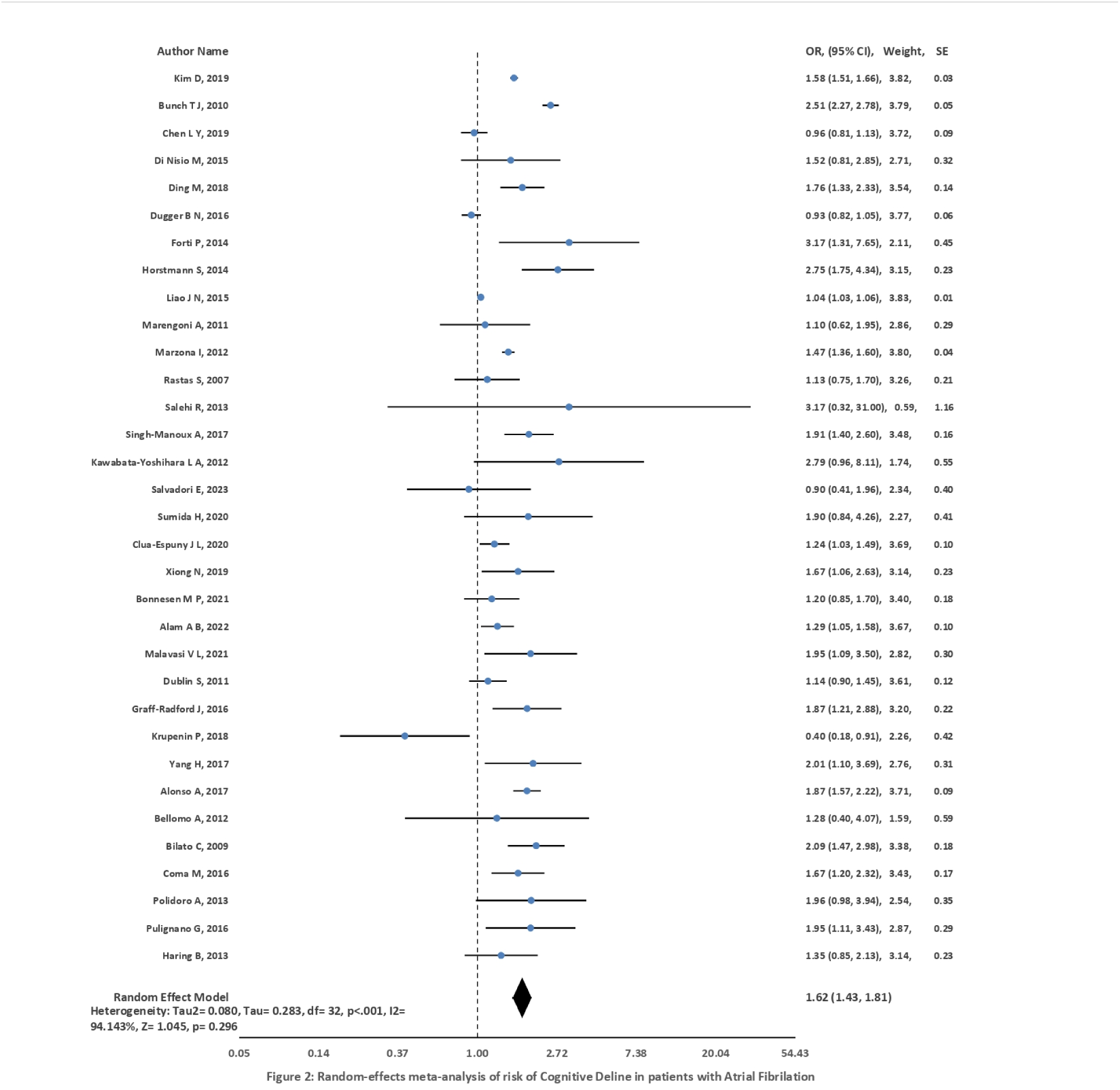
Random Effects Meta Analysis Of Risk Of Cognitive Decline In Atrial Fibrillation Patients

**Figure 3:**
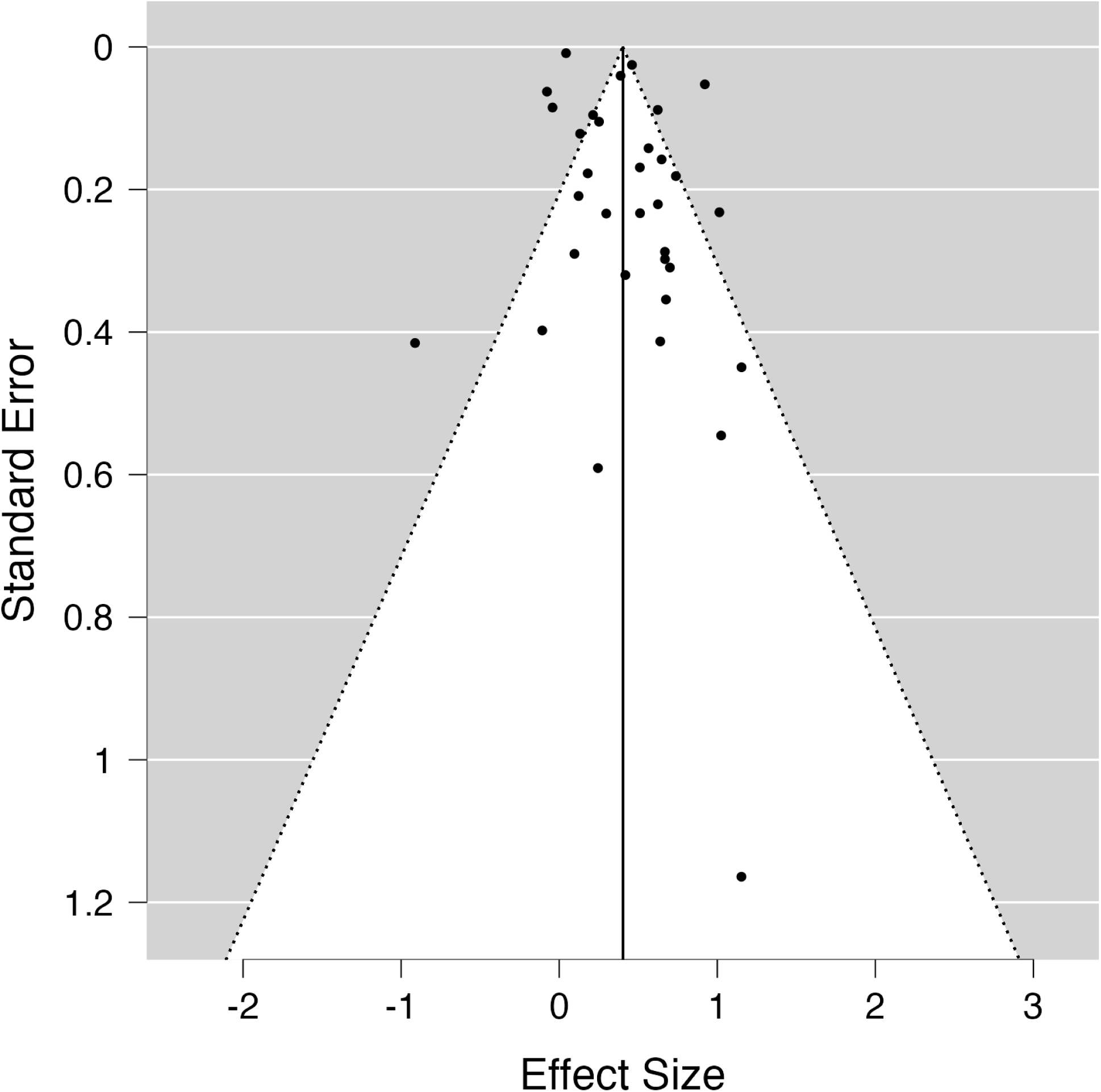
Funnel Plot

#### Hypertension

A pooled analysis of 19 studies^12,18,22,23,24,27,28,30,32,34,37–39,43,45,46,49–51^ that included patients with concomitant hypertension revealed a significant correlation between AF and CImp (OR: 1.73; 95% CI: 1.49, 1.97; df: 18). Hypertension was a covariate that had a significant degree of between-study heterogeneity (91.42%). However, it was unable to fully account for the variation in effect size (Q = 130.887; df, 17; p =0.001).

#### Diabetes Mellitus

A substantial correlation between AF and CImp was found in the pooled analysis of 20 studies^12,18,22,23,24,27,28,30,32,34,37,38,39,43,45,48–51^ that included individuals with DM comorbidity (OR: 1.75; 95% CI: 1.52, 1.98; df: 19). Similar to hypertension, DM as a covariate exhibited substantial between-study heterogeneity (92.52%) and did not fully explain the variation in effect size (Q = 219.471; df = 18; p < 0.001).

#### Hyperlipidemia

The pooled analysis of the seven studies^18,27,28,32,43,49,50^ that included patients with hyperlipidemia as a comorbidity revealed a significant correlation between AF and CImp (OR: 2.00; 95% CI: 1.50, 2.51; df: 6). When utilizing hyperlipidemia as a covariate, the between-study heterogeneity was considerable (44.645%), but it could account for all of the variations in the effect magnitude (Q = 7.168; df = 5; p = 0.179).

#### Myocardial Infarction (MI)

A substantial correlation was found between AF and CImp (OR: 1.75; 95% CI: 1.44, 2.06; df: 7), according to a pooled analysis of eight studies^12,18,23,30,37,48,49,50^ reporting patients with MI comorbidity. When MI was included as a covariate, there was significant between-study heterogeneity (90.334%), but it did not fully explain the observed variation in effect size (Q = 85.425; df = 6; p< 0.001).

#### Heart Failure (HF)

The pooled analysis showed a significant correlation between AF and CImp among 10 studies^12,22,23,24,28,37,39,46,48,50^ reporting patients with HF comorbidity (OR: 1.49; 95% CI: 1.24, 1.74; df: 9). When utilizing HF as a covariate, there was significant between-study heterogeneity (90.765%), and it could not completely account for the observed variability in effect size (Q = 148.580; df = 8; p< 0.001).

#### Stroke

The pooled analysis of the 10 studies^22,30,38,39,43,46,48,49,50,51^ that reported patients with concomitant strokes revealed a significant correlation between AF and CImp (OR: 1.63; 95% CI: 1.39, 1.88; df: 9). When stroke was included as a covariate, there was considerable between-study heterogeneity (75.695%), but it was unable to fully account for the variation in effect size (Q = 38.029; df = 8; p< 0.001).

#### Smoking

The pooled analysis of the eleven studies^12,18,23,24,30,38,40,43,46,49,50^ that included patients who smoked showed a substantial correlation between AF and CImp (OR: 1.74; 95% CI: 1.52, 1.96; df: 10). When smoking was included as a covariate, there was considerable between-study heterogeneity (41.625%), and the smoking covariate could account for all of the variation in effect size (Q = 11.196; df = 9; p = 0.263).

### Sensitivity Analysis using Sub-groups

#### Study Design

Even when the analysis was restricted to certain study designs, significant findings were still observed for the combined endpoint of CImp and/or dementia. There was a significant correlation in 23 prospective studies^12,22,24,26,29–31,33,35,36,38,39,41,42,43,44,46–52^ (OR: 1.57; 95% CI: 1.38, 1.75; df: 22; I^2^ = 79.278%). A significant correlation was also seen in cross-sectional studies, which included 4 studies^25,27,34,45^ (OR: 2.06; 95% CI: 1.16, 2.96; df: 3; I^2^ = 84.313%).

Case-control studies (2 studies^23,32^) (OR: 1.96; 95% CI: 0.53, 3.39; df: 1) and retrospective studies (4 studies^18,28,37,40^) (OR: 1.52; 95% CI: 0.86, 2.18; df: 3; I^2^ = 98.199%) resulted in non-significant association.

### Comprehensive Cognitive Testing

Sub-groups that used more sensitive neuropsychological tests^12,18,22,26–29,31,33–40,42,44–46^ rather than relying simply on MMSE testing, showed that AF was substantially associated with an increased risk of CImp (OR: 1.60, 95% CI: 1.31, 1.88, df: 19, I^2^ = 96.367%). Similarly, AF was shown to be substantially linked with CImp in subgroups that solely underwent MMSE testing^23,24,30,41,47–52^ (OR: 1.70, 95% CI: 1.52, 1.87, df: 9, I^2^ = 53.896%).

## DISCUSSION

This systematic review and meta-analysis aimed to assess the relationship between AF and CImp. The analysis of 33 observational studies revealed a significant association between AF and an elevated risk of the combined endpoint of CImp and/or dementia. Additional analyses were conducted, focusing on studies that employed sensitive neuropsychological testing, to address the heterogeneity in methodologies used to identify CImp and these secondary analyses further supported the substantial correlation between AF and CImp. The observed variability can be attributed to variations in sample size, study design and population differences.

To address the issue of significant heterogeneity, we used a random-effects model and several subgroups, sensitivity, and meta-regression analyses. However, still significant heterogeneity existed. Importantly, our study exclusively included individuals without a history of stroke, revealing a strong association between AF and CImp.

The study by Kokkinidis et. al. revealed that individuals with AF have 3.11 times higher odds of developing dementia, 1.60 times higher odds of CImp, and a 2.26 times higher odds of experiencing either CImp/dementia compared to those without AF^54^. Several other studies also found similar associations between AF and CImp^9,11,33,47,55–58^. However, further investigation is needed to understand how AF affects cognitive performance in diverse ethnic and geographic populations. Interestingly, sex and race have been found to interact significantly with CImp among individuals with AF^59^.

In addition to its risk of thromboembolism, recent research suggests that AF increases the chance of CImp through various pathophysiological pathways and processes. Silent brain infarctions, microemboli, microbleeds, brain atrophy, cerebral hypoperfusion, altered hemostatic function, vascular oxidative stress, and inflammation are a few reasons that contribute to this association^8^. These factors may lead to more severe CImp in patients with chronic or permanent AF, longer duration of AF, or a higher burden of AF^60^. Furthermore, AF is associated with an inflammatory state, and it is unknown whether AF-induced systemic inflammation contributes to dementia^9^. Both CImp and AF share several common risk factors, as they primarily affect the elderly population. Silent brain infarctions, and clinically apparent ischemic strokes are likely processes behind the elevated risk of CImp complicating AF in this age group^60,61^. Some studies suggest that anticoagulant therapy, including direct oral anticoagulants and vitamin K antagonists, may reduce the incidence of CImp and dementia in patients with AF. However, silent brain infarction still occurs among individuals with AF despite on anticoagulation^62^. Therefore, randomized trials^56^ are still needed to provide conclusive evidence that anticoagulation reduces the risk of CImp and dementia in AF patients. Future research should also explore the potential advantages of increasing the use of oral anticoagulation or enhancing adherence to anticoagulant treatment in specific patient subgroups^61,63^.

AF has been associated with decreased volume in the right basal frontal lobe and right inferior cerebellum^64^, taking into account the existence of hypertension and a history of heart failure. There is growing evidence that the aging process, hypertension, and AF are all connected to structural and functional changes in various parts of the central nervous system that affect autonomic control^65^. Furthermore, the link between AF and dementia persists even after accounting for common risk factors such as diabetes and hypertension^9^. According to Bano et. al.^66^, diabetes is associated with a higher likelihood of neurological comorbidities such as stroke (OR: 1.39; 95% CI: 1.03, 1.87) and CImp (OR: 1.75; 95% CI: 1.39, 2.21). Therefore, effective management of co-existent risk factors such as hypertension, diabetes, and so on and how it will impact the association of AF with CImp should be explored in large clinical trials.

Several limitations should be considered when interpreting the findings of this study. The inclusion of case-control and retrospective studies, along with prospective studies led to greater heterogeneity and therefore limits our ability to draw any firm conclusions. This highlights the need for further research using robust study designs to elucidate the relationship between AF and CImp. The included studies employed diverse methods and criteria to identify AF, CImp, and dementia which likely contributed to significant heterogeneity. We addressed this by employing a random-effects model and carrying out numerous subgroup, sensitivity, and meta-regression studies. The studies had a wide confidence interval, which suggested differences in the research methodology and participant demographics may have contributed to less accurate estimates. Although the exclusion of outlier studies led to a modest decrease in heterogeneity, it did not reach statistical significance.

In conclusion, this meta-analysis provides strong evidence linking the association between AF and poor cognitive function. Despite between-study heterogeneity and the influence of multiple comorbidities, the findings demonstrate a link between AF and an increased risk of CImp. Future studies should focus on investigating the underlying reasons and potential therapeutic interventions to mitigate cognitive decline in AF patients. Given high morbidity and its significant impact on the quality of life, our findings have implications for patients and healthcare systems considering the burden of AF due to increasing life expectancy.

1. Contributors: Study design, literature review, statistical analysis: AA. Data management, drafting manuscript: AA, MAM, MIA. Access to data: AA, MAM, MIA, EZS Manuscript revision, intellectual revisions, mentorship: MIA, EZS Final approval: AA, MAM, MIA, EZS
2. Funders: No funds, grants, or other support was received.
3. Prior presentations: This paper has not been presented anywhere else.

## Data Availability

The original contributions presented in the study are included in the article/Supplementary Material, further inquiries can be directed to the corresponding author/s.

## List of Abbreviations

AF: Atrial Fibrillation
CImp: Cognitive Impairment
MMSE: Mini Mental State Exam
MoCA: Montreal Cognitive Assessment

